# Gemykibivirus detection in acute encephalitis patients from Nepal

**DOI:** 10.1101/2024.02.13.24302648

**Authors:** Eans Tara Tuladhar, Smita Shrestha, Susan Vernon, Lindsay Droit, Kathie A. Mihindukulasuriya, Mamta Tamang, Lata Karki, Annie Elong Ngono, Bimlesh Jha, Bal Krishna Awal, Bimal Sharma Chalise, Runa Jha, Sujan Shresta, David Wang, Krishna Das Manandhar

## Abstract

Acute Encephalitis Syndrome (AES) causes significant morbidity and mortality worldwide. In Nepal, Japanese encephalitis virus (JEV) accounts for ∼ 5-20% of AES cases, but ∼75% of AES cases are of unknown etiology. We identified a gemykibivirus in CSF collected in 2020 from a male child with AES using metagenomic next-generation sequencing. Gemykibiviruses are single stranded, circular DNA viruses in the family *Genomoviridae*. The complete genome of 2211 nucleotides was sequenced which shared 98.69% nucleotide identity to its closest relative, Human associated gemykibivirus 2 isolate SAfia-449D. Two real-time PCR assays were designed, and screening of 337 CSF and 164 serum samples from AES patients in Nepal collected in 2020 and 2022 yielded 11 CSF and 1 serum sample that were positive in both PCR assays. Complete genomes of 7 of the positives were sequenced. These results identify a candidate etiologic agent of encephalitis in Nepal.

## Introduction

Encephalitis is a neurological disorder associated with a high mortality rate on a global scale (*1*). It is the inflammation of the brain parenchyma with clinical features of fever, altered mental state, and/or new onset of seizures. At present, low sociodemographic index regions in Asia and Africa carry the highest burden of encephalitis (*2*). In the year 2021, a total of 512 cases of AES were reported in Nepal (*3*). More than 100 different infectious agents that cause encephalitis are known including bacteria, viruses, fungi, and parasites (*4, 5*). The major known etiologic agents of encephalitis as reported in Nepal and internationally are Japanese encephalitis virus (JEV), enteroviruses, herpes simplex, and varicella zoster viruses (*6, 7*). In Nepal, there has been a national surveillance program for JE since 2004 wherein CSF and serum samples of suspected viral cases of encephalitis are collected from sentinel sites throughout Nepal with technical support from WHO and analyzed at the National Public Health Laboratory (NPHL) by serology for anti-JEV IgM. In numerous studies in Nepal assessing cases since 2000, ∼70-95% of the AES cases per year have no diagnosis (*3, 8-11*). A significant fraction of encephalitis in other countries similarly lack diagnosis, despite extensive testing (*5, 12, 13*). In recent years, the application of metagenomic analysis to patients with encephalitis has begun to identify a range of emerging viruses linked to encephalitis (*14-19*) The *Genomoviridae* family of viruses has single stranded DNA genomes of ∼2.1-2.2 kb (*20*) that encode a capsid protein (CP) and a replication associated protein (REP). They have been identified from a wide range of hosts including plants, insects, animals, and humans (*20-22*). There are ten genera in the *Genomiviridae* family (*20*). Viruses in the genus *Gemykibivirus* have been identified in multiple human cases and in multiple specimen types including: blood of febrile Tanzanian children (*23*); the respiratory tract of an elderly woman with respiratory distress in China (*24*); feces from diarrhea patients in Brazil (*25*); blood of healthy blood donors of Brazil (*21*); blood of HIV positive Cameroonian males (*26*); and in cervical swab of HIV/HPV infected pregnant females (*27*). Specific to encephalitis, there are reports of gemykibiviruses in CSF from an encephalitic child from China (*15*) and CSF from three patients with encephalitis from Sri Lanka (*25*). Furthermore, analysis of Nepalese sewage yielded the complete genome of a gemykibivirus (*25*). Here, we used metagenomic next generation sequencing (NGS) to identify the presence of a gemykibivirus in CSF from a patient with encephalitis in Nepal. Further PCR screening identified 12 additional positive cases.

## Materials and Methods

### Ethical clearance, study population, and collection of biospecimens

This study was approved by the Nepal Health Research Council of Nepal [approval# 274-2020] and the Human Research Protection Office of Washington University in Saint Louis [202004087]. The study population focused on AES patients who were negative for JE IgM. Residual samples from national JE surveillance sentinel sites in Nepal were utilized. The index case CSF sample in the NPHL repository was collected in 2020 from Rupandehi district. For prevalence studies, 122 repository specimens (82 CSF, 40 sera) collected in 2020 and 379 repository specimens (255 CSF, 124 sera) from 2022 were tested by PCR for gemykibivirus.

### Total nucleic acid extraction

Total nucleic acid extraction was performed using the Invitrogen Pure Link ™ Viral RNA/DNA mini kit [Thermo Fisher Scientific] and eluted in 50 μL volume following the instruction manual of the kit. Samples were stored at -80°C.

### Metagenomic NGS analysis

Extracted total nucleic acid were randomly amplified as described previously (*28*) and used for library construction with NEBNext Ultra DNA Library Prep Kit for Illumina (New England Biolabs). The sample library was sequenced on the Illumina MiSeq instrument using the 2 × 250 bp paired-end protocol. NGS data was analyzed for presence of viruses using CZID (*29*). NGS data is available at ENA: PRJEB72279.

### Genome sequencing of index case and additional cases

The NGS contigs were confirmed using PCR, cloning, and sanger sequencing with primers (Appendix Table 1). Using a pair of primers that amplified the entire circular genome, Gemy1xgenomeF (5’TTAATCGATCTAGAGGATCCTTGTTAGATATCCATATGGCGG-3’) and Gemy1xgenomeR (5’-TTAGTAATGGGCCCGGATCCACGAGAGGAACACG-3’), three independent PCR reactions were performed, and the resulting fragments were cloned into pCR4.0 and sequenced using the Oxford Nanopore technologies (Plasmidsauraus). Additional positive cases were similarly amplified and whole genomes sequenced to 3X coverage. Complete genomes sequences are available at Genbank (Accession# PP270194-PP270201).

**Table 1:** Patient demographics and qPCR Ct. values of gemykibivirus positive cases.

| Sample ID<sup>†</sup> | Specimen | Age group<br>[yrs] | Gender | Ct.<br>Gemy_1<sup>‡</sup> | Ct.<br>Gemy_2<sup>§</sup> | Specimen<br>collection date | Districts<sup>¶</sup> | Province |
| --- | --- | --- | --- | --- | --- | --- | --- | --- |
| N0000218 | CSF | 11-15 | M | 32.42 | 30.68 | 2020 Feb | Kaski | Gandaki |
| N0000256 | CSF | 0-5 | F | 29.41 | 29.34 | 2020 July | Myagdi | Gandaki |
| N0000300 | CSF | 6-10 | F | 32.95 | 31.46 | 2020 Sep | Palpa | Lumbini |
| N0000358 | Serum | 0-5 | M | 29.27 | 29.94 | 2022 Feb | Sarlahi | Madhesh |
| N0000359 | CSF | 0-5 | F | 27.4 | 27.08 | 2022 Feb | Kapilvastu | Lumbini |
| N0000395 | CSF | 0-5 | M | 30.74 | 30.73 | 2022 April | Kaski | Gandaki |
| N0000434 | CSF | 56-60 | M | 29.02 | 30.06 | 2022 June | Lalitpur | Bagmati |
| N0000468 | CSF | 30-35 | F | 30.84 | 31.47 | 2022 July | Chitwan | Bagmati |
| N0000490 | CSF | unknown | M | 29.86 | 32.08 | 2022 July | Syangja | Gandaki |
| N0000545 | CSF | 70-75 | F | 26.84 | 25.35 | 2022 Aug | Kathmandu | Bagmati |
| N0000546 | CSF | 20-25 | M | 24.65 | 23.53 | 2022 Aug | Kathmandu | Bagmati |
| N0000722 | CSF | 20-25 | F | 28.74 | 29.72 | 2022 Nov | Chitwan | Bagmati |

### Gemykibivirus qPCR development

Two sets of Taqman real time PCR primers and probes were designed using express software (Applied Biosystems), one targeting the CP gene and the other targeting the REP gene. The primers and the probes were supplied by IDT (Integrated DNA Technologies, USA). The first assay, Gemy_1, targets the CP gene using primers GemykibiTM_8917F (5’-ACCTCTTATCCGGTTTGGCA-3’) and GemykibiTM_8917R (5’-AGCGCGAAATTCCTCTTGAC-3’) and the probe GemykibiTM_8917Probe(5’- [6-FAM]CGGACCTGA[ZEN]CCGGATGCCCGG[3IABkFQ]-3’) that uses FAM and the dual quencher Zen and Iowa Black. The second assay, Gemy_2, targets the REP gene with GemykibiTM_9967F (5’-GGTCAGAGCCTAGTGTTGTATG-3’) GemykibiTM_9967R (5’-CGACGTTGTCTGTGTCTTCT-3’) GemykibiTM_9967Probe (5’-[6-FAM]AAGACACTC[ZEN]TGGGCAAGAAGCC TT[IABkFQ]-3’) using the same fluor and quencher.

For both assays, standard curves were generated using serial 10-fold dilution ranging from 2x10^8^ to 2x10^1^ copies of positive control plasmid [plasmid PCR4 containing the respective target sequence]. A 20 μL PCR mixture was made comprising 2 μL of extracted nucleic acid sample, 10 μL of 2x TaqMan Fast Advanced Master Mix [Thermo Fisher Scientific], and 5 pmol of each primer and probe. The PCR reactions were performed in 96-well plates on a CFX Opus 96 thermocycler [Bio-Rad] with one negative control nuclease free water in each row and one positive control of 2 x10^3^ copies per plate. The cycling conditions were 50°C for 2 mins, 95°C for 30 secs and 40 cycles of 95°C for 5 secs followed by 60°C for 30 secs. The threshold of all plates was set at standard value and data was analyzed using Bio-Rad CFX Maestro 2.3 software. Samples were counted as positive if their threshold cycle (Ct) value was less than 33.

### Phylogenetic analysis

Representative protein sequences of the REP gene of prototypes of each genus in the *Genomoviridae* were downloaded from GenBank. Alignments were generated using Clustal Omega (*30*). The alignment converted to fasta via http://sequenceconversion.bugaco.com/converter/biology/sequences/clustal_to_fasta.php. Maximum likelihood trees were generated with bootstrapping, using W-IQ-TREE (*31*). Trees were visualized using iTOL (*32*). All available complete genomes in the species *Gemykibivirus humas2* were downloaded from Genbank along with representative genomes from the species *Gemykibivirus humas 1, 3, 4*, and *5*, and the top 10 additional complete genomes with highest BLASTn scores. Alignments were generated using Clustal Omega (*30*) Maximum likelihood trees were generated with bootstrapping, using W-IQ-TREE (*31*).Trees were visualized using iTOL (*32*).

## Results

### Detection of a gemykibivirus by metagenomic NGS

NGS of nucleic acids extracted from the CSF of a male child yielded reads that could be assembled into two contigs that shared 97 and 99% nucleotide similarity with human associated Gemykibivirus2 SAfia-449D (accession# MN765187.1). Using PCR, gaps between the two contigs were spanned to generate a complete circular genome of 2211 nt. To formally assess the taxonomic relationship of this virus to viruses in the family *Genomoviridae*, we generated a maximum likelihood phylogenetic tree of the REP protein with the type species of each genus, in accordance with the ICTV guidelines (*20, 22*) (Figure 1A). The virus was most similar to the prototype virus from the genus *Gemykibivirus*. To further assess its relationship within the genus *Gemykibivirus*, we generated a maximum likelihood tree using the whole genome sequence (Figure 1B), which demonstrated it is most closely related to Human gemykibivirus 2 SAfia-449D, a virus detected in blood of Tanzanian children (*23*), with 98.69% nucleotide identity. Based on these criteria, the virus genome from the index case was designated Human gemykibivirus 2 Nepal/N0000051/2020. Human gemykibivirus 2 Nepal/N0000051/2020 was also closely related to Gemycircularvirus-SL1 (accession# KP133075), a previously reported gemykibivirus detected in CSF from an encephalitis patient in Sri Lanka (*25*) sharing 97.96% nucleotide identity (33 SNPs across the genome and a 12 bp insertion located in a region of tandem repeat hexamers) and it shared 98.64% identity to another gemykibivirus, Gemycircularvirus NP (accession# KP133080), detected in sewage from Nepal (*25*).

**Figure 1.**
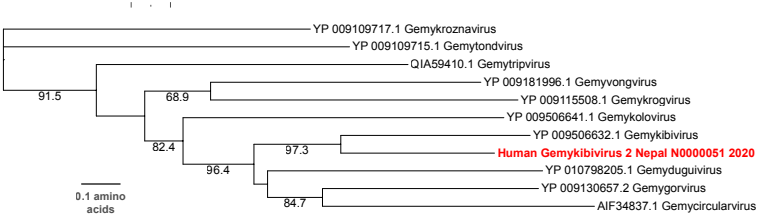
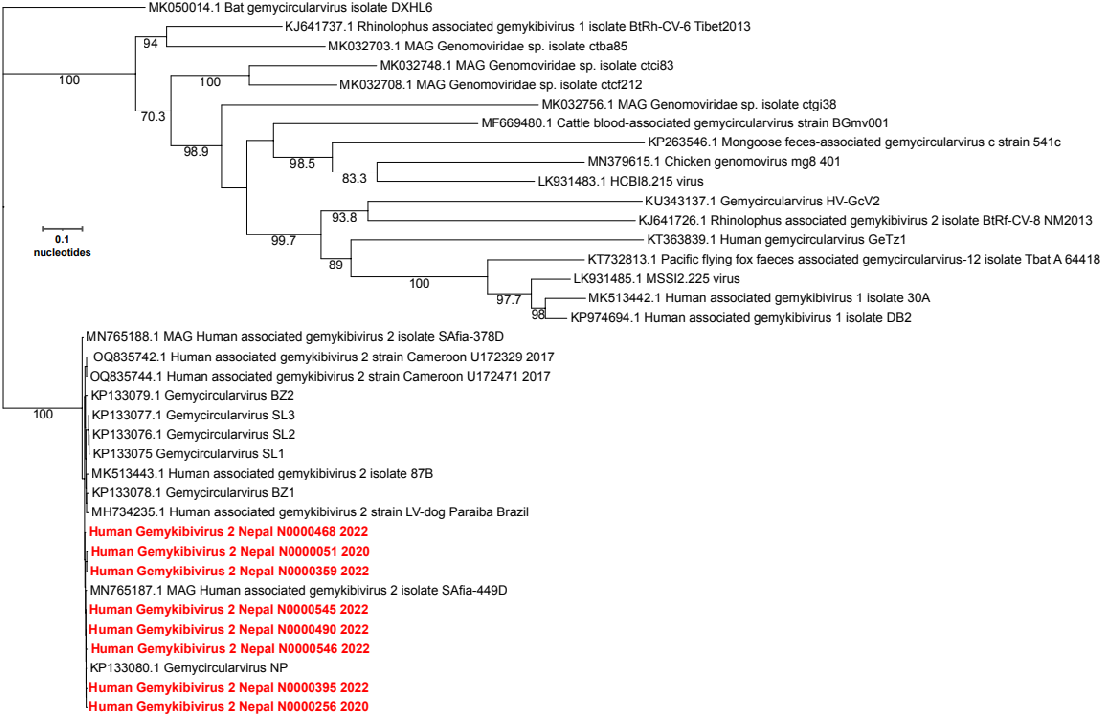
Phylogenetic trees of (A) amino acid sequences of the index case REP protein and representatives of each genus in the family *Genomoviridae;* (B) whole genome nucleotide sequences of the positive samples from Nepal compared to other genomes within the genus *Gemykibivirus*.

Gene predictions identified three ORFs, characteristic of Gemykibiviruses (Figure 2). The CP ORF is 969 bp, the REP ORF, generated by splicing is 1114 bp, and the unknown ORF3, which overlaps with the REP ORF, is 702 bp. A large intergenic region (LIR) of 127bp is present, and the putative viral origin of replication nona-nucleotide motif 5′-TAAAATTTA-3′ described in Gemycircularvirus NP (accession# KP133080) is conserved. A predicted stem loop in the LIR is observed from nucleotides 23 to 57. The stem loop structure is present in genomoviruses and geminiviruses where it is necessary for rolling circle replication (*33, 34*).

**Figure 2.**
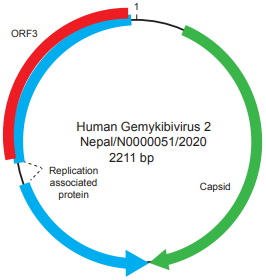
Schematic of the genome of Human gemykibivirus 2 Nepal/N0000051/2020.

### Identification of additional positive specimens from encephalitis patients

To define the prevalence of this virus, two Taqman real-time PCR assays were designed and validated, one targeting the REP gene and one targeting the CP gene (Appendix Figure 1). A total of 337 CSF samples and 164 serum samples were tested using both assays. Sample with Ct. values <33 for both assays were considered positive. There were three gemykibivirus positive CSF specimens from 2020 and 9 positive specimens (8 CSF and 1 serum) from 2022 (Table). The prevalence rate in CSF was 3.3% (11/337) and in serum was 0.6% (1/164). Positive patients ranged from 4 months to 72 years of age. Including the index case, there were 7 male and 6 female patients. Geographically, most positive patients were from districts in south-central Nepal (Figure 3). Unfortunately, no additional clinical details are available for the patients in this study besides meeting the acute case definition of encephalitis.

**Figure 3.**
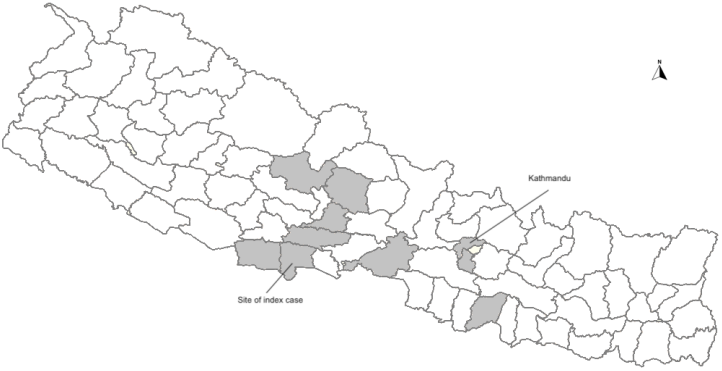
Map of Nepal and locations of the gemykibivirus positive cases [shaded].

### Whole genome sequencing and phylogenetic analysis of additional positive cases

From the 10 positive specimens with highest Gemykibivirus copy number, we tried to amplify the whole genome using PCR. For 7 of the samples, we obtained amplicons that corresponded to the whole genome. These were cloned and sequenced to 3X coverage to determine their consensus sequences. These 7 genomes varied from the index genome by 4 to 21 SNPs. The intron of the Rep gene contains a region with seven tandemly repeated hexamers, and deletions of one or more of the hexamers are observed in some of the genomes. Phylogenetic analysis of the complete genomes demonstrated that they formed a clade that included known Gemykibiviruses previously detected in human CSF, stool, and blood.

## Discussion

We used metagenomic NGS to detect Human gemykibivirus2 Nepal/N0000051/2020 in CSF of a male encephalitic child from Rupandehi district (Figure 3) of Nepal which is alongside the border with India. Further screening through qPCR identified an additional 12 positive specimens from patient samples collected in 2020 and 2022. We found that these cases were mostly concentrated in and around the south-central region of Nepal (Figure 3).

The identified virus genomes were very closely related (97.96% nt. identity) to a gemykibivirus previously detected in CSF from three encephalitis patients from Sri Lanka (*25*). In addition, a distinct gemykibivirus (Human gemykibivirus 4) has also been reported in CSF of an encephalitis patient from China (*15*). Together with our study, these data implicate viruses in the genus *Gemykibivirus* as potential causal agents of encephalitis. There is one report of *Gemykibivirus* in sewage from Nepal from 2012, which is also highly similar to the sequences we detected in Nepalese patients from 2020-2022. This suggests that gemykibivirus has been circulating in Nepal for at least the past decade. Furthermore, detection in sewage, raises the possibility that gemykibivirus may be transmitted fecal-orally, similarly to some neurotropic viruses such as polio and enteroviruses. Detection of highly similar viruses in patients in Tanzania (*23*) and Brazil (*21*) suggests that gemykibiviruses are globally widespread.

One limitation of this study is that the samples analyzed were residual specimens from a surveillance repository without additional available clinical metadata, thus limiting our knowledge of the precise symptoms, disease severity, and outcomes of these patients. While detection of gemykibiviruses in presumptively sterile CSF supports the hypothesis that they could be causal agents of encephalitis, additional research to culture the virus and establish animal models to fulfill Koch’s postulates are needed to definitively establish causality. In addition, more prevalence studies in encephalitis and other diseases are also needed as are serological studies to define the extent of human infection by gemykibiviruses. Finally, additional, unbiased approaches are needed to define the etiologies of encephalitis in Nepal, and worldwide.

## Data Availability

All data produced in the present work are contained in the manuscript

## Acknowledgment

This project was supported by National Institutes of Health grant U01AI151810.

## Biographical sketch of first author

Dr. Tuladhar, MD is currently pursuing a PhD at Tribhuvan University Central Department of Biotechnology, Nepal studying emerging viral infections.

### Appendices

Appendix Table: Primers used for PCR

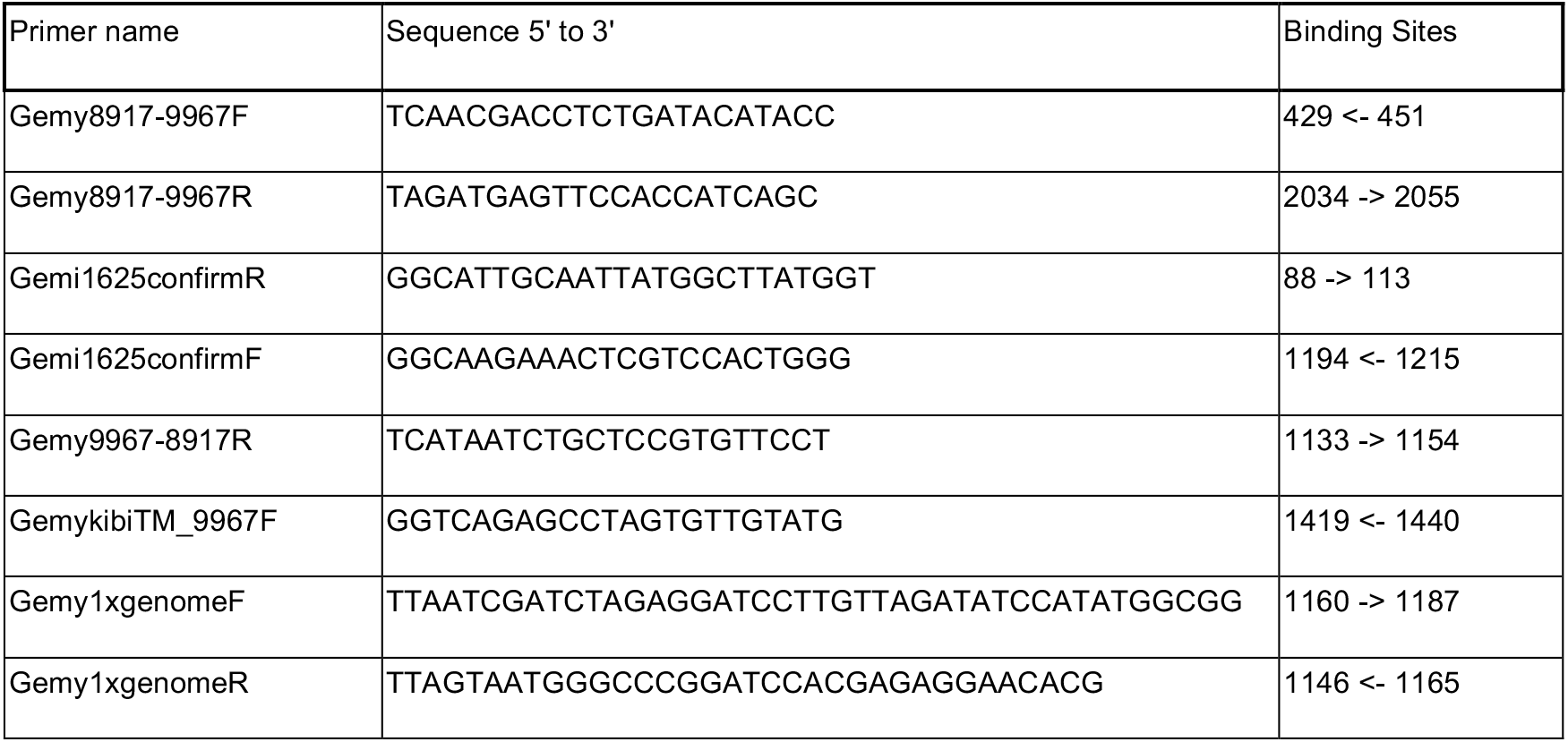

## Notes

### Competing Interest Statement

The authors have declared no competing interest.

### Funding Statement

This project was supported by National Institutes of Health grantU01AI151810

### Author Declarations

Ethics committee of Nepal Health Research Council of Nepal gave ethical approval for this work[approval# 274-2020] and Ethics committee of the Human Research Protection Office of Washington University in Saint Louis gave ethical approval for this work[202004087].

